# Factors associated with Low birth weight in Cambodia: A secondary analysis of Cambodia Demographic and Health Survey, 2021-2022

**DOI:** 10.1101/2024.02.25.24303351

**Authors:** Samnang Um, Sok Sakha, Leng Dany

**Affiliations:** National Institute of Public Health, Phnom Penh, Cambodia; Elite Angkor Clinic, Siem Reap, Cambodia

**Keywords:** Low birth weight, Newborn, Cambodia Demographic and Health Survey

## Abstract

Low birth weight (LBW) is a serious global health concern, including in Cambodia. It is associated with a higher risk of neonatal, infant, and under-five mortality and morbidity. We examined the prevalence of LBW across provinces and factors associated with LBW among newborns in Cambodia. We analyzed the most recent children’s data from the Cambodia Demographic and Health Survey (CDHS) 2021-2022. There were 4,565 weighted newborns included in this study. The provincial variation in the prevalence of LBW was done using ArcGIS. Multiple logistic regression analyses were conducted to assess factors independently associated with LBW. The prevalence of LBW was 5.9% (95% CI: 5.2–6.8), which was highest in Ratanak Kiri (13.9%), Kratie (12.8%), Pailin (12.7%), and Svay Rieng (10.2%). Factors were independently associated with LBW, including mothers with primary education (AOR = 1.82; 95% CI: 1.09–3.04), unemployed (AOR = 1.62; 95% CI: 1.03–2.55), being a first-born child (AOR = 1.74; 95% CI: 1.24–2.45), born to mothers in rural areas (AOR = 1.42; 95% CI: 1.00–2.01). However, mothers who attended at least 4 ANC visits had lower odds of LBW (AOR = 0.65; 95% CI: 0.47–0.91). The prevalence of LBW among newborns in Cambodia is lower than in Southsea Asia and worldwide. However, provincial variations in the prevalence of LBW were high in remote areas. Public Health programs should target provinces where LBW prevalence remains high. Cambodia policy continues to recommend four or more ANC visits to pregnant women, particularly those who have limited education and live in rural areas.

## Introduction

Newborn low birth weight (LBW) is defined as a weight at birth of less than 2500 grams (or 5.5 pounds) by the World Health Organization (WHO) [1]. LBW is a serious public health issue affecting developed and developing nations [1]. Globally, in 2020, an estimated 19.8 million (or 14.6%) live births were suffering from LBW [1,2]. According to estimates, LBW is more common in South Asia (28%), East Asia and the Pacific (6%), sub-Saharan Africa (13%), and Latin America (9%) [3,4]. Compared to East Asia and the Pacific as a whole, Cambodia has a higher proportion of LBW. World Bank data shows that 11% of newborns were LBW in 2000. After that, prevalence declined to around 8% of all live births in 2005 [5] and has remained stable at about 8% in 2010 and 7.9 in 2014 but slightly declined to about 5.9% in 2021-22 [5–7]. However, newborns with LBW are more common in rural areas (7.2%) than in urban areas (4%). The highest rates of LBW were seen in remote and low-income provinces like Ratanakiri (14.8%), Pailin (12.9%), Mondolkiri (10.7%), and Stung Treng (10.2%) in 2021–22 [7]. Approximately 2.04% of all deaths were related to LBW in Cambodia [8]. Furthermore, compared to newborns with normal birth weight, those with LBW had over five times increased risk of dying before turning five years old [9]. Children’s mortality and morbidity are independently associated with newborns with LBW [10,11]. Also, it increases the risk for chronic non-communicable diseases (NCDs), such as diabetes and cardiovascular disease, in adulthood [1,12,13]. Lack of iron folic acid (IFA) supplementation and deworming during pregnancy[14], maternal weight gain [15], preterm birth [16], mothers having inadequate antenatal care visits [17], anemic [18], underweight mothers during pregnancy [19], use of tobacco [20], and alcohol consumption during pregnancy [21] were associated with increased risk of LBW. Therefore, the risk of LBW is 3.8 times higher for mothers who are under 20 years or over 35 years compared to mothers who are between the ages of 20-35 years [22]. In general, mothers of low socio-economic status commonly have LBW babies. For example, mothers from the poorest (AOR = 1.7) and middle-income households (AOR = 1.6) were more likely to deliver LBW babies than the richest mothers [23]. A previous study in Cambodia indicated that factors independently associated with increased odds of LBW included the mother’s no education with an adjusted odds ratio (AOR = 1.6), babies born to mothers with less than four antenatal care (ANC) visits during the pregnancy (AOR = 2.0) [17].

The Royal Government of Cambodia (RGC) has recognized that nutrition is the highest priority program in Cambodia and has been closely monitored by the Cambodian Prime Minister to enforce policy implementation to reach the Sustainable Development Goal [24]. Cambodia aims to reduce LBW to less than 6% in 2023 and 4% in 2025 [24]. To reduce LBW, the government has taken different strategies such as counseling during ANC visits, routine supplementation of IFA to pregnant women, providing nutritious food such as super cereals to pregnant women residing in highly food insecure areas, and awareness of early pregnancy, smoking, and alcohol during pregnancy [14]. Despite these efforts, the prevalence of LBW remains a significant public health problem in Cambodia. The proportion of LBW in Cambodia has slowly decreased in newborns during 2010–2022, particularly among remote provinces and vulnerable mothers [5–7]. In addition, LBW has been documented as an independent factor associated with childhood malnutrition [1,2,25], infectious disease [1,4,26], and risk of under-five mortality in Cambodia [1,4,9]. To the authors’ knowledge, a limited study investigated the association between socio-economic and maternal factors and LBW using updated data, which has not been explored. A prior study described the prevalence of LBW across provinces in Cambodia, and its associated factors have been utilized since 2014 [17]. Therefore, this study aimed to examine the prevalence of LBW across provinces and factors associated with LBW among newborns in Cambodia using updated CDHS 2021-22 data. Hence, this study would help better understand the maternal, child, and household socio-economic factors of LBW among newborns in Cambodia and may help develop policies and programs with more effective strategies and interventions to reduce the prevalence of LBW, thereby contributing further to the reduction of newborn mortality in Cambodia.

## Material and Methods

### Ethics Statement

The CDHS survey was approved by the National Ethical Committee for Health Research, Ministry of Health of Cambodia on May 10, 2021 (**Ref: 083 NECHR**), and the Institutional Review Board (IRB) of ICF in Rockville, Maryland, USA. Which are publicly accessible upon request through the DHS website at (URL: https://dhsprogram.com/data/available-datasets.cfm) [27]. Before conducting the interview, field researchers explained the objective of the survey to the parents/guardians of each participant under 18 years of age, and written informed consent was obtained from their parents/guardians.

### Data sources

We followed the methods of Chhea et al. 2018 [17]. To analyze the prevalence of LBW across provinces and factors associated with LBW among newborns in Cambodia. This study was a retrospective secondary data analysis using children’s data (KR file) from the most recent CDHS 2021–22, a nationally representative population-based household survey from September 15, 2021, to February 15, 2022. To select participants from every province, CDHS employed a two-stage stratified cluster sampling technique [7]. In the first stage, 709 clusters, or enumeration areas (EAs), were selected (241 EAs from urban areas and 468 EAs in rural areas). In the second stage, a systematic random sample was applied between 25-30 households from each cluster or EA for a total sample size of 21,270 families. Interviews were then conducted with 19,496 women aged 15– 49 who had birth five years before the survey, with a response rate of 98.2%. About 8,153 children’s information was collected from their mothers [7]. Our analysis was restricted to the last pregnancy with the most recent live births two years preceding the survey and reported birth weight. As a result, we excluded 3,295 not-last births and 187 newborns with missing birth weight data. A final sample included in this analysis was 4,671 newborns (4,565 weighted counts).

### Measurements Outcome variable

The birth weight of the newborn was the outcome variable. LBW is defined as a weight at birth of less than 2500 grams [1]. Then, the original variable was recoded into the binary variable, where non-LBW= 0 (if birth weight ≥ 2,500 grams) and LBW= 1 (if birth weight < 2,500 grams).

### Independent variables

**Maternal characteristics included**: Women’s age in years (≤ 19, 20-29, ≥ 30), marital status (married, not married), and education (no education, primary, and secondary/higher). Employment status (not working, non-professional, and professional). Having the first ANC visit in the first trimester of pregnancy (yes, no), the number of ANC visits (less than 4 visits, 4 or more visits), having ANC provided by skilled personnel such as a medical doctor, nurse, or midwife (yes, no), receiving a tetanus injection (not, one dose, two or more), giving iron tablets during pregnancy, and intestinal parasite medication (yes, no), and having adequate ANC components was defined six interventions including blood pressure measurement, blood sample collected, urine sample collected, health and nutrition education, ultrasound conducted, and counseling on danger signs. Types of delivered (normal birth, cesarean birth), and place of delivered (home, health facilities). Maternal smoking cigarettes (non-smoker vs smoker) and alcohol drinking (no, yes). Healthcare barriers (no barrier, one or more possible barriers reported included distance, money, and waiting time), and health insurance coverage (yes, no). **Child characteristics** included sex of child (male, female), birth interval (first child, < 2 years, 2-3 years, and 3+), and birth order (1, 2–3, 4+). **Socio-economic characteristics** included households’ wealth index (poorest, poorer, middle, richer, richest). The wealth index is calculated via principal component analysis (PCA) using variables for household assets and dwelling characteristics [7], type of toilet facility (improved, unimproved), and source of drinking water (improved, unimproved). Place of residence (rural, urban). Cambodia’s provinces were regrouped for analytic purposes into a categorical variable with four geographical regions: plains, Tonle Sap, coastal/sea, and mountains.

### Statistical Analysis

Statistical analyses were performed using STATA version 18 (Stata Crop 2021, College Station, TX). We formally incorporated the DHS’s complex sample design using the “survey” package; all estimations were carried out using the **svy** command in our descriptive and logistic regression analyses. Descriptive statistics for the maternal, child, and household socio-economics characteristics were described using weighted frequency distributions. The provincial variation in the prevalence of LBW was done using ArcGIS software version 10.8 [28]. A shapefile for Cambodian administrative boundaries was obtained from the United Nations for Coordination of Humanitarian Affairs (OCHA) (URL: https://data.humdata.org/dataset/cod-ab-khm?). License (URL: https://data.humdata.org/faqs/licenses). Bivariate analysis using Chi-square tests assessed associations between independent variables (maternal, child, and household socio-economics characteristics) and LBW. The final multiple logistic regression analyses included variables associated with LBW at p-value ≤ 0.10 [25] or that had a potential confounder variable, including maternal age at the time of giving birth. Simple logistic regression determined the magnitude of associations between LBW and maternal, child, and household socio-economic characteristics. Results are reported as odds ratios (OR) with 95% confidence intervals (CI). Multiple logistics regression was then used to assess independent factors associated with LBW after adjusting for other potential confounding factors in the model. Results from the final adjusted model are reported as adjusted odds ratios (AOR) with 95% CI and corresponding p-values. Multicollinearity between independent variables was checked before fitting the final regression model, including maternal age at the time of giving birth, educational level, employment status, number of ANC visits during pregnancy, child’s birth order, wealth index, and place of residence. The result of evaluating variance inflation factor (VIF) scores after fitting the regression model with the mean = 1.20 indicated no collinearity concerns [29].

## Results

### Characteristics of the study samples

**Table 1** describes the household socio-demographic characteristics of the 4,565 last births newborns with the most recent live births in the two years preceding the survey, including their reported birth weight and the characteristics of their mothers. Nearly 70% of the mothers were age 20–29 at the time of childbirth, and 11% of the mothers had no schooling, 40.4% had a primary education, 49.1% had a secondary or higher education, 31% had no working, and about 95% of mothers were married. Only 1.1% of the mothers smoked cigarettes, while 36.3% reported drinking alcohol in the past few months. Most mothers (86.3%) attended at least four ANC visits during pregnancy. Likewise, 87.8% initiated ANC within the first trimester of pregnancy, and 98.7% attended with a skilled provider. The majority (89.1%) of mothers consumed iron supplements, and more than eight-ten (82.3%) of the mothers took parasite medication during pregnancy. About 60.7% of the mothers received adequate ANC components. Approximately 24.8% of mothers have health insurance. Newborns delivered at the hospital were (96.8%). Over half of the newborns were boys; singleton births were 99.5%. Among the sample, 99.1%% live births. Their average weight at birth was 3,088 grams. 32.4% of newborns are 1st order births, and 10.2% are 4^th^ or high-order births. About 40.2% of newborns were born into low-wealth index households, and 60.8% were born in rural areas (see **Table 1).**

**Table 1.** Descriptive statistics of the latest children, mothers, and household socio-demographic characteristics were included in the study, CDHS 2021-2022 (N = 4,565 weighted count).

| Variables | Total Births | % |
| --- | --- | --- |
| **Maternal Characteristics** |  |  |
| **Age at the time of giving birth** |  |  |
| ≤ 19 | 1,083 | 23.7 |
| 20-29 | 3,145 | 68.9 |
| ≥ 30 | 338 | 7.4 |
| **Educational level** |  |  |
| No education | 484 | 10.6 |
| Primary | 1,842 | 40.4 |
| Secondary/higher | 2,240 | 49.1 |
| **Employment status** |  |  |
| Not working | 1395 | 30.6 |
| Professional | 1273 | 27.9 |
| Non-professional | 1897 | 41.6 |
| **Marital status** |  |  |
| Not married | 231 | 5.1 |
| Married | 4,334 | 94.9 |
| **Smoking cigarettes** |  |  |
| Non-Smoker | 4,515 | 98.9 |
| Smoker | 50 | 1.1 |
| **Alcohol intake in the past month** |  |  |
| No | 2,910 | 63.7 |
| Yes | 1,655 | 36.3 |
| **Number of ANC visits during pregnancy** |  |  |
| < 4 visits | 625 | 13.7 |
| ≥ 4 visits | 3,940 | 86.3 |
| **First ANC visit in the first trimester** |  |  |
| No | 501 | 11.0 |
| Yes | 4,007 | 87.8 |
| **Antenatal care from a skilled provider** |  |  |
| Unskilled | 61 | 1.3 |
| Skilled | 4,504 | 98.7 |
| **Tetanus toxoid injections during pregnancy** |  |  |
| None | 767 | 16.8 |
| One dose | 666 | 14.6 |
| Two or more | 3,133 | 68.6 |
| **Iron supplementation during pregnancy** |  |  |
| No | 499 | 10.9 |
| Yes | 4,067 | 89.1 |
| **Parasite medication during pregnancy** |  |  |
| No | 806 | 17.7 |
| Yes | 3,759 | 82.3 |
| **Compliance with ANC components** |  | 0.0 |
| Inadequate | 1,797 | 39.4 |
| Adequate | 2,769 | 60.7 |
| **Healthcare Barriers** |  |  |
| No barrier | 1,869 | 40.9 |
| One or More Barriers | 2,696 | 59.1 |
| **Health insurance coverage** |  |  |
| No | 3,432 | 75.2 |
| Yes | 1,133 | 24.8 |
| **Child's characteristics** |  |  |
| **Place of birth** |  |  |
| Home | 147 | 3.2 |
| Health facilities | 4,418 | 96.8 |
| **Sex of child** |  |  |
| Female | 2,233 | 48.9 |
| Male | 2,332 | 51.1 |
| Single birth | 4,544 | 99.5 |
| Alive after birth | 4,525 | 99.1 |
| Average birth weight (SD) | 3,088 grams (SD = 9.8) |  |
| Max-Min | 500 - 5500 grams |  |
| **Child's birth interval (year)** |  |  |
| First child | 1,479 | 32.4 |
| < 2 years | 305 | 6.7 |
| 2-3 years | 500 | 11.0 |
| Three or more | 2,281 | 50.0 |
| **Child's birth order** |  |  |
| 1st child | 1,479 | 32.4 |
| 2-3 child | 2,619 | 57.4 |
| Four or more | 467 | 10.2 |
| **Type of delivery** |  |  |
| Normal Delivery | 3,717 | 81.4 |
| Cesarean Section | 848 | 18.6 |
| **Household characteristics** |  |  |
| **Wealth index** |  |  |
| Poor | 1,835 | 40.2 |
| Middle | 847 | 18.6 |
| Rich | 1,883 | 41.2 |
| **Type of toilet facility** |  |  |
| Improved | 4,000 | 87.6 |
| Non-Improved | 565 | 12.4 |
| **Source of drinking water** |  |  |
| Improved | 2,747 | 60.2 |
| Unimproved | 1,818 | 39.8 |
| **Place of residence** |  |  |
| Urban | 1,789 | 39.2 |
| Rural | 2,776 | 60.8 |
| **Geographic Region** |  |  |
| Plain | 2,239 | 49.0 |
| Tonle Sap | 1,408 | 30.8 |
| Coastal | 270 | 5.9 |
| Plateau/Mountain | 649 | 14.2 |

**Notes:** Survey weights are applied to obtain weighted percentages. \***Plains:** Phnom Penh, Kampong Cham, Tbong Khmum, Kandal, Prey Veng, Svay Rieng, and Takeo; **Tonle Sap:** Banteay Meanchey, Kampong Chhnang, Kampong Thom, Pursat, Siem Reap, Battambang, Pailin, and Otdar Meanchey; **Coastal/sea:** Kampot, Kep, Preah Sihanouk, and Koh Kong; **Mountains:** Kampong Speu, Kratie, Preah Vihear, Stung Treng, Mondul Kiri, and Ratanak Kiri.

### Distribution of newborns with LBW by provinces

The prevalence of low birth weight was 5.9% (95% CI: 5.2–6.8). And low birth weight is highest among newborn in Ratanak Kiri (13.9%), Kratie (12.8%), Pailin (12.7%), Svay Rieng (10.2%), Stung Treng (9.2%), Tboung Khmum (8.3%) and lowest among newborn in Kampong Speu (1.5%), Banteay Meanchey (2.5%) and Pursat (2.6%), and Kampong Chhnang (2.7%) (See **Fig 1**, and **S1 Table)**.

**Fig 1.**
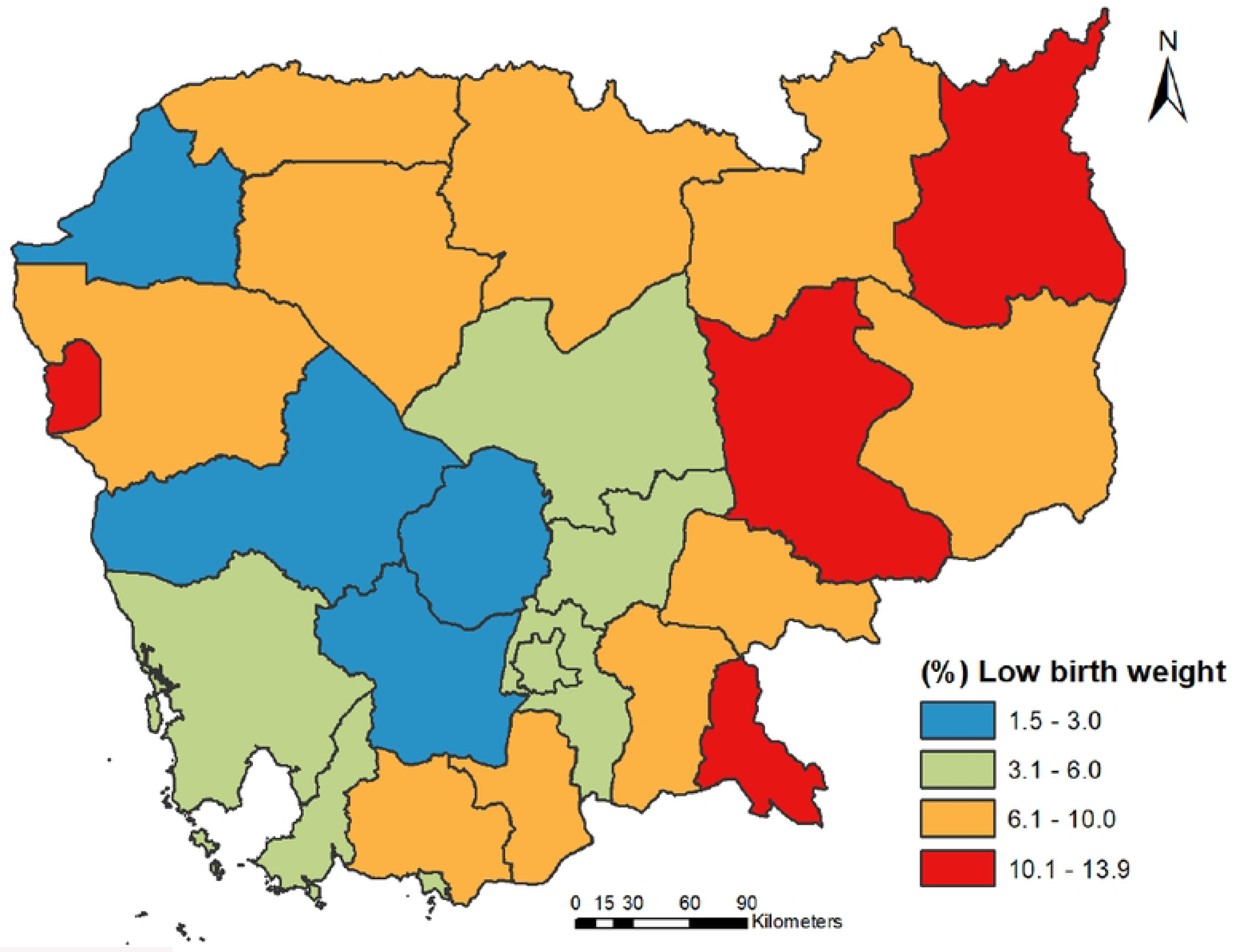
Prevalence of newborns with low birth weight by province. The map was created using ArcGIS software version 10.8 [28]. A shapefile for Cambodian administrative boundaries was obtained from the United Nations for Coordination of Humanitarian Affairs (OCHA) (https://data.humdata.org/dataset/cod-ab-khm<u>?).</u> License (https://data.humdata.org/faqs/licenses).

### Factors associated with LBW among the most recent live births

In bivariate Chi-square test presents the prevalence of LBW by independent variables, household and mother’s characteristics and behaviors, access to health care during pregnancy, health insurance coverage, maternal health status, and child characteristics (see Table 2). Newborns had a higher prevalence of LBW if they were born to mothers who had completed primary education (4.4%) compared to higher education (p-value = 0.076), were unemployed job (7.4%) compared to other careers, or were from poor households (7.1%) compared to rich households (p-value = 0.046), and born to rural area had a higher prevalence of LBW than those in urban areas (6.9% vs 4.57%, p-value = 0.005). In addition, LBW prevalence among children born to mothers attending less than 4 ANC visits during pregnancy (8.6%) was significantly higher than for those born to mothers attending four or more ANC visits (5.5%) (p-value = 0.007). First-born babies had a higher LBW prevalence (8%) than second- and third-born babies (4.8%) and fourth-born or higher (6.4%) (p-value = 0.004). There were no differences in the prevalence of LBW between boy and girl babies, place and types of delivered, mother’s age, marital status, smoking status, alcohol consumption, initial ANC in the first trimester, consumed iron supplement, took parasite medication, adequate ANC components during pregnancy, perceptions of problems in accessing health service, and health insurance coverage (see **Table 2**).

**Table 2.**
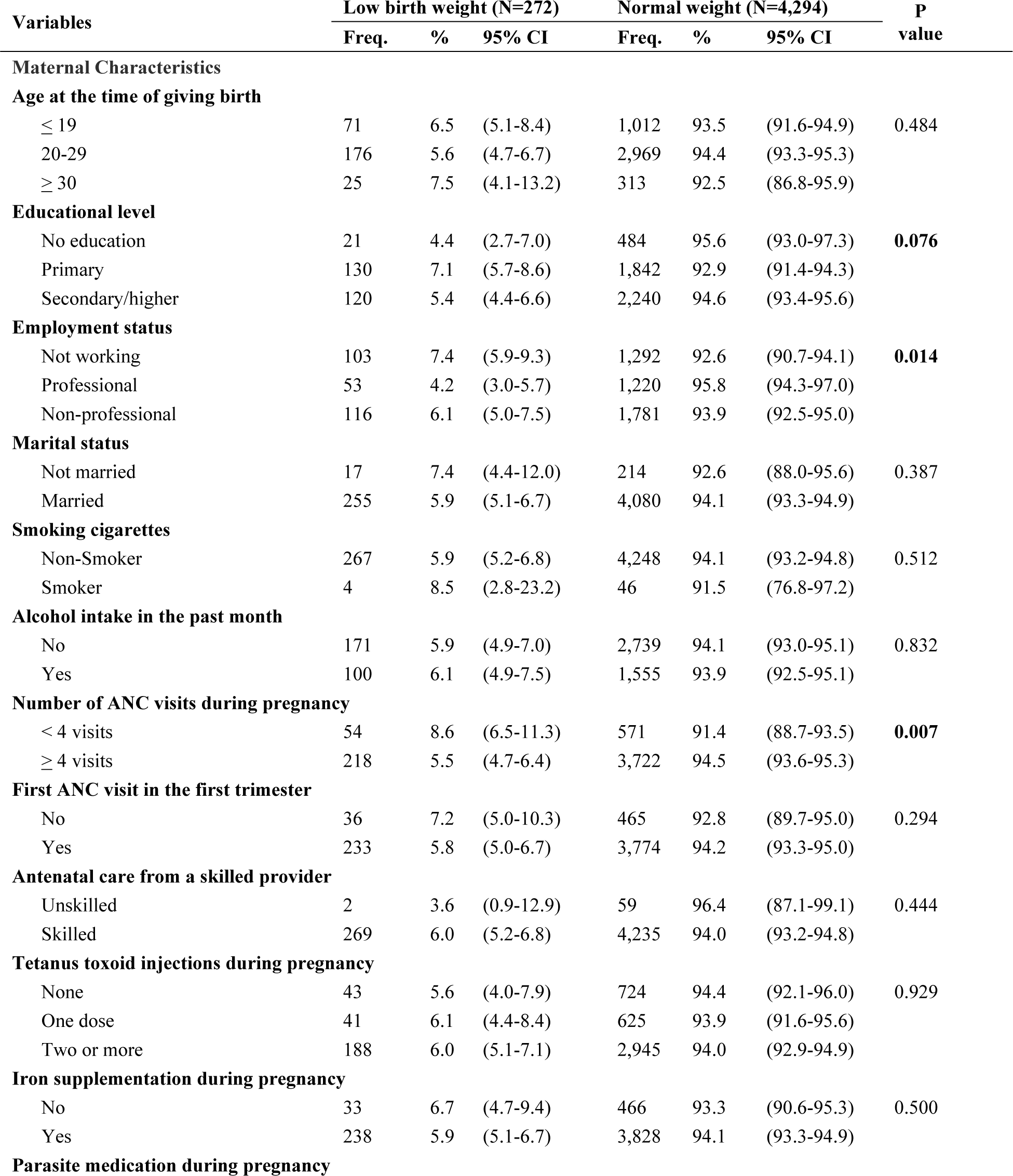
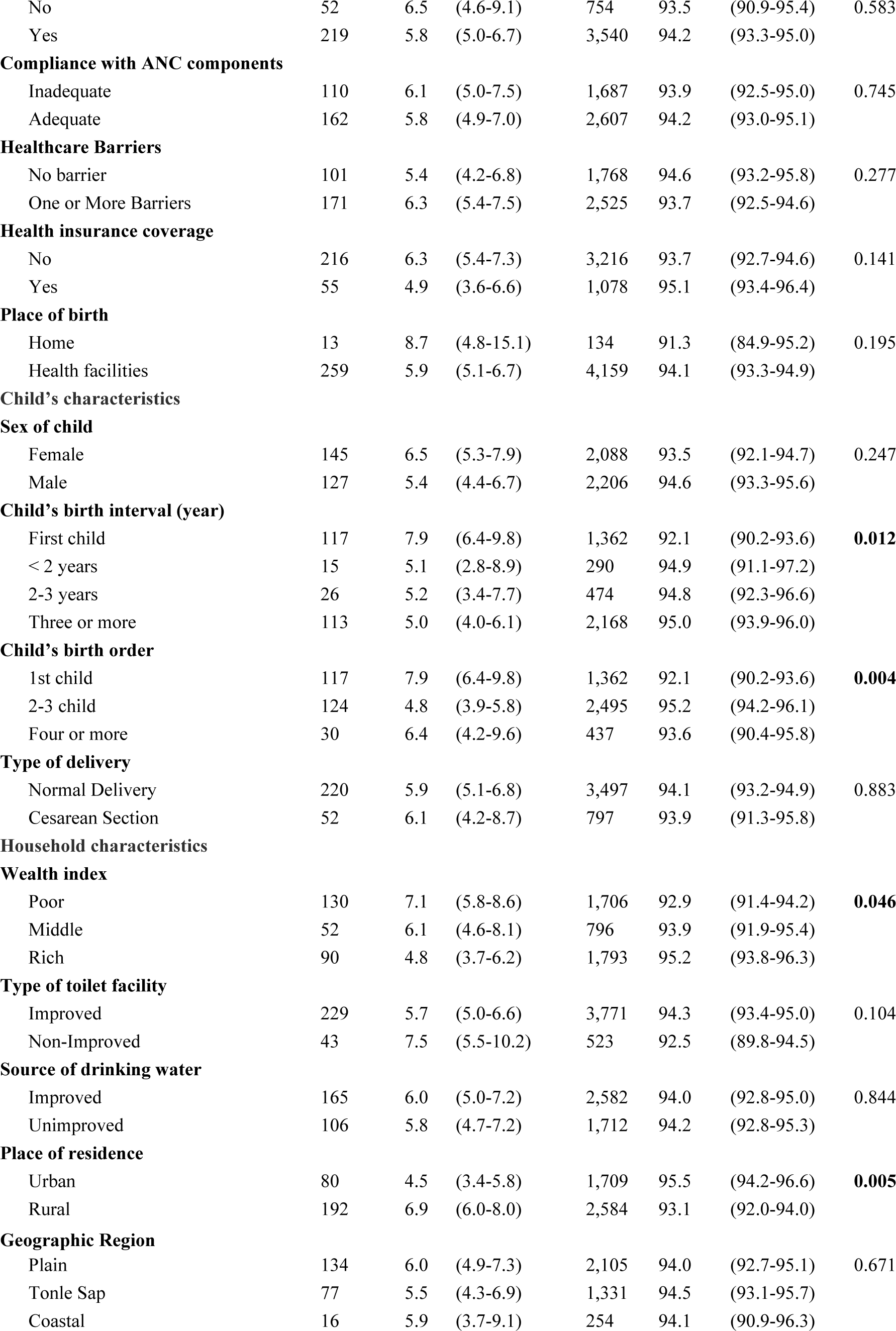
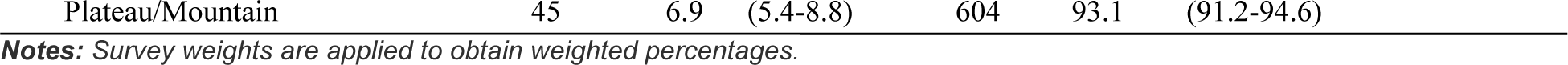
Distribution of socio-demographic factors and proportion of newborns with LBW in Cambodia, 2021-2022 (N = 4,565, weighted count).

| Variables | Low birth weight (N=272) |  |  | Normal weight (N=4,294) |  |  | P value |
| --- | --- | --- | --- | --- | --- | --- | --- |
|  | Freq. | % | 95% CI | Freq. | % | 95% CI |  |
| **Maternal Characteristics** |  |  |  |  |  |  |  |
| **Age at the time of giving birth** |  |  |  |  |  |  |  |
| ≤ 19 | 71 | 6.5 | (5.1-8.4) | 1,012 | 93.5 | (91.6-94.9) | 0.484 |
| 20-29 | 176 | 5.6 | (4.7-6.7) | 2,969 | 94.4 | (93.3-95.3) |  |
| ≥ 30 | 25 | 7.5 | (4.1-13.2) | 313 | 92.5 | (86.8-95.9) |  |
| **Educational level** |  |  |  |  |  |  |  |
| No education | 21 | 4.4 | (2.7-7.0) | 484 | 95.6 | (93.0-97.3) | **0.076** |
| Primary | 130 | 7.1 | (5.7-8.6) | 1,842 | 92.9 | (91.4-94.3) |  |
| Secondary/higher | 120 | 5.4 | (4.4-6.6) | 2,240 | 94.6 | (93.4-95.6) |  |
| **Employment status** |  |  |  |  |  |  |  |
| Not working | 103 | 7.4 | (5.9-9.3) | 1,292 | 92.6 | (90.7-94.1) | **0.014** |
| Professional | 53 | 4.2 | (3.0-5.7) | 1,220 | 95.8 | (94.3-97.0) |  |
| Non-professional | 116 | 6.1 | (5.0-7.5) | 1,781 | 93.9 | (92.5-95.0) |  |
| **Marital status** |  |  |  |  |  |  |  |
| Not married | 17 | 7.4 | (4.4-12.0) | 214 | 92.6 | (88.0-95.6) | 0.387 |
| Married | 255 | 5.9 | (5.1-6.7) | 4,080 | 94.1 | (93.3-94.9) |  |
| **Smoking cigarettes** |  |  |  |  |  |  |  |
| Non-Smoker | 267 | 5.9 | (5.2-6.8) | 4,248 | 94.1 | (93.2-94.8) | 0.512 |
| Smoker | 4 | 8.5 | (2.8-23.2) | 46 | 91.5 | (76.8-97.2) |  |
| **Alcohol intake in the past month** |  |  |  |  |  |  |  |
| No | 171 | 5.9 | (4.9-7.0) | 2,739 | 94.1 | (93.0-95.1) | 0.832 |
| Yes | 100 | 6.1 | (4.9-7.5) | 1,555 | 93.9 | (92.5-95.1) |  |
| **Number of ANC visits during pregnancy** |  |  |  |  |  |  |  |
| < 4 visits | 54 | 8.6 | (6.5-11.3) | 571 | 91.4 | (88.7-93.5) | **0.007** |
| ≥ 4 visits | 218 | 5.5 | (4.7-6.4) | 3,722 | 94.5 | (93.6-95.3) |  |
| **First ANC visit in the first trimester** |  |  |  |  |  |  |  |
| No | 36 | 7.2 | (5.0-10.3) | 465 | 92.8 | (89.7-95.0) | 0.294 |
| Yes | 233 | 5.8 | (5.0-6.7) | 3,774 | 94.2 | (93.3-95.0) |  |
| **Antenatal care from a skilled provider** |  |  |  |  |  |  |  |
| Unskilled | 2 | 3.6 | (0.9-12.9) | 59 | 96.4 | (87.1-99.1) | 0.444 |
| Skilled | 269 | 6.0 | (5.2-6.8) | 4,235 | 94.0 | (93.2-94.8) |  |
| **Tetanus toxoid injections during pregnancy** |  |  |  |  |  |  |  |
| None | 43 | 5.6 | (4.0-7.9) | 724 | 94.4 | (92.1-96.0) | 0.929 |
| One dose | 41 | 6.1 | (4.4-8.4) | 625 | 93.9 | (91.6-95.6) |  |
| Two or more | 188 | 6.0 | (5.1-7.1) | 2,945 | 94.0 | (92.9-94.9) |  |
| **Iron supplementation during pregnancy** |  |  |  |  |  |  |  |
| No | 33 | 6.7 | (4.7-9.4) | 466 | 93.3 | (90.6-95.3) | 0.500 |
| Yes | 238 | 5.9 | (5.1-6.7) | 3,828 | 94.1 | (93.3-94.9) |  |
| **Parasite medication during pregnancy** |  |  |  |  |  |  |  |
| No | 52 | 6.5 | (4.6-9.1) | 754 | 93.5 | (90.9-95.4) | 0.583 |
| Yes | 219 | 5.8 | (5.0-6.7) | 3,540 | 94.2 | (93.3-95.0) |  |
| Compliance with ANC components |  |  |  |  |  |  |  |
| Inadequate | 110 | 6.1 | (5.0-7.5) | 1,687 | 93.9 | (92.5-95.0) | 0.745 |
| Adequate | 162 | 5.8 | (4.9-7.0) | 2,607 | 94.2 | (93.0-95.1) |  |
| Healthcare Barriers |  |  |  |  |  |  |  |
| No barrier | 101 | 5.4 | (4.2-6.8) | 1,768 | 94.6 | (93.2-95.8) | 0.277 |
| One or More Barriers | 171 | 6.3 | (5.4-7.5) | 2,525 | 93.7 | (92.5-94.6) |  |
| Health insurance coverage |  |  |  |  |  |  |  |
| No | 216 | 6.3 | (5.4-7.3) | 3,216 | 93.7 | (92.7-94.6) | 0.141 |
| Yes | 55 | 4.9 | (3.6-6.6) | 1,078 | 95.1 | (93.4-96.4) |  |
| Place of birth |  |  |  |  |  |  |  |
| Home | 13 | 8.7 | (4.8-15.1) | 134 | 91.3 | (84.9-95.2) | 0.195 |
| Health facilities | 259 | 5.9 | (5.1-6.7) | 4,159 | 94.1 | (93.3-94.9) |  |
| Child's characteristics |  |  |  |  |  |  |  |
| Sex of child |  |  |  |  |  |  |  |
| Female | 145 | 6.5 | (5.3-7.9) | 2,088 | 93.5 | (92.1-94.7) | 0.247 |
| Male | 127 | 5.4 | (4.4-6.7) | 2,206 | 94.6 | (93.3-95.6) |  |
| Child's birth interval (year) |  |  |  |  |  |  |  |
| First child | 117 | 7.9 | (6.4-9.8) | 1,362 | 92.1 | (90.2-93.6) | 0.012 |
| < 2 years | 15 | 5.1 | (2.8-8.9) | 290 | 94.9 | (91.1-97.2) |  |
| 2-3 years | 26 | 5.2 | (3.4-7.7) | 474 | 94.8 | (92.3-96.6) |  |
| Three or more | 113 | 5.0 | (4.0-6.1) | 2,168 | 95.0 | (93.9-96.0) |  |
| Child's birth order |  |  |  |  |  |  |  |
| 1st child | 117 | 7.9 | (6.4-9.8) | 1,362 | 92.1 | (90.2-93.6) | 0.004 |
| 2-3 child | 124 | 4.8 | (3.9-5.8) | 2,495 | 95.2 | (94.2-96.1) |  |
| Four or more | 30 | 6.4 | (4.2-9.6) | 437 | 93.6 | (90.4-95.8) |  |
| Type of delivery |  |  |  |  |  |  |  |
| Normal Delivery | 220 | 5.9 | (5.1-6.8) | 3,497 | 94.1 | (93.2-94.9) | 0.883 |
| Cesarean Section | 52 | 6.1 | (4.2-8.7) | 797 | 93.9 | (91.3-95.8) |  |
| Household characteristics |  |  |  |  |  |  |  |
| Wealth index |  |  |  |  |  |  |  |
| Poor | 130 | 7.1 | (5.8-8.6) | 1,706 | 92.9 | (91.4-94.2) | 0.046 |
| Middle | 52 | 6.1 | (4.6-8.1) | 796 | 93.9 | (91.9-95.4) |  |
| Rich | 90 | 4.8 | (3.7-6.2) | 1,793 | 95.2 | (93.8-96.3) |  |
| Type of toilet facility |  |  |  |  |  |  |  |
| Improved | 229 | 5.7 | (5.0-6.6) | 3,771 | 94.3 | (93.4-95.0) | 0.104 |
| Non-Improved | 43 | 7.5 | (5.5-10.2) | 523 | 92.5 | (89.8-94.5) |  |
| Source of drinking water |  |  |  |  |  |  |  |
| Improved | 165 | 6.0 | (5.0-7.2) | 2,582 | 94.0 | (92.8-95.0) | 0.844 |
| Unimproved | 106 | 5.8 | (4.7-7.2) | 1,712 | 94.2 | (92.8-95.3) |  |
| Place of residence |  |  |  |  |  |  |  |
| Urban | 80 | 4.5 | (3.4-5.8) | 1,709 | 95.5 | (94.2-96.6) | 0.005 |
| Rural | 192 | 6.9 | (6.0-8.0) | 2,584 | 93.1 | (92.0-94.0) |  |
| Geographic Region |  |  |  |  |  |  |  |
| Plain | 134 | 6.0 | (4.9-7.3) | 2,105 | 94.0 | (92.7-95.1) | 0.671 |
| Tonle Sap | 77 | 5.5 | (4.3-6.9) | 1,331 | 94.5 | (93.1-95.7) |  |
| Coastal | 16 | 5.9 | (3.7-9.1) | 254 | 94.1 | (90.9-96.3) |  |

|  |  |  |  |  |  |  |
| --- | --- | --- | --- | --- | --- | --- |
| Plateau/Mountain | 45 | 6.9 | (5.4-8.8) | 604 | 93.1 | (91.2-94.6) |

*Notes: Survey weights are applied to obtain weighted percentages.*

### Factors associated with LBW among the most recent live births in multiple logistic regression

As shown in **Table 3**, several factors were independently associated with increased odds of being LBW among newborns with the most recent live births in the two years preceding the survey. These factors included children born to mothers with primary education (AOR = 1.82; 95% CI: 1.09–3.04) and unemployed mothers (AOR = 1.62; 95% CI: 1.03–2.55). Being a first-born child was still strongly associated with LBW (AOR = 1.74; 95% CI: 1.24–2.45) compared with being the second child. Children born to mothers in rural areas had 1.42 times higher odds of LBW than those born to urban mothers (AOR = 1.42; 95% CI: 1.00–2.01). However, mothers who attended at least 4 ANC visits during their last pregnancy had 0.65 times lower odds of LBW compared with women having less than 4 ANC visits (AOR = 0.65; 95% CI: 0.47–0.91). We did not find that the effects of mothers’ age and household wealth index were statistically insignificant after adjusting for other confounding factors in the multiple logistic regression model.

**Table 3.**
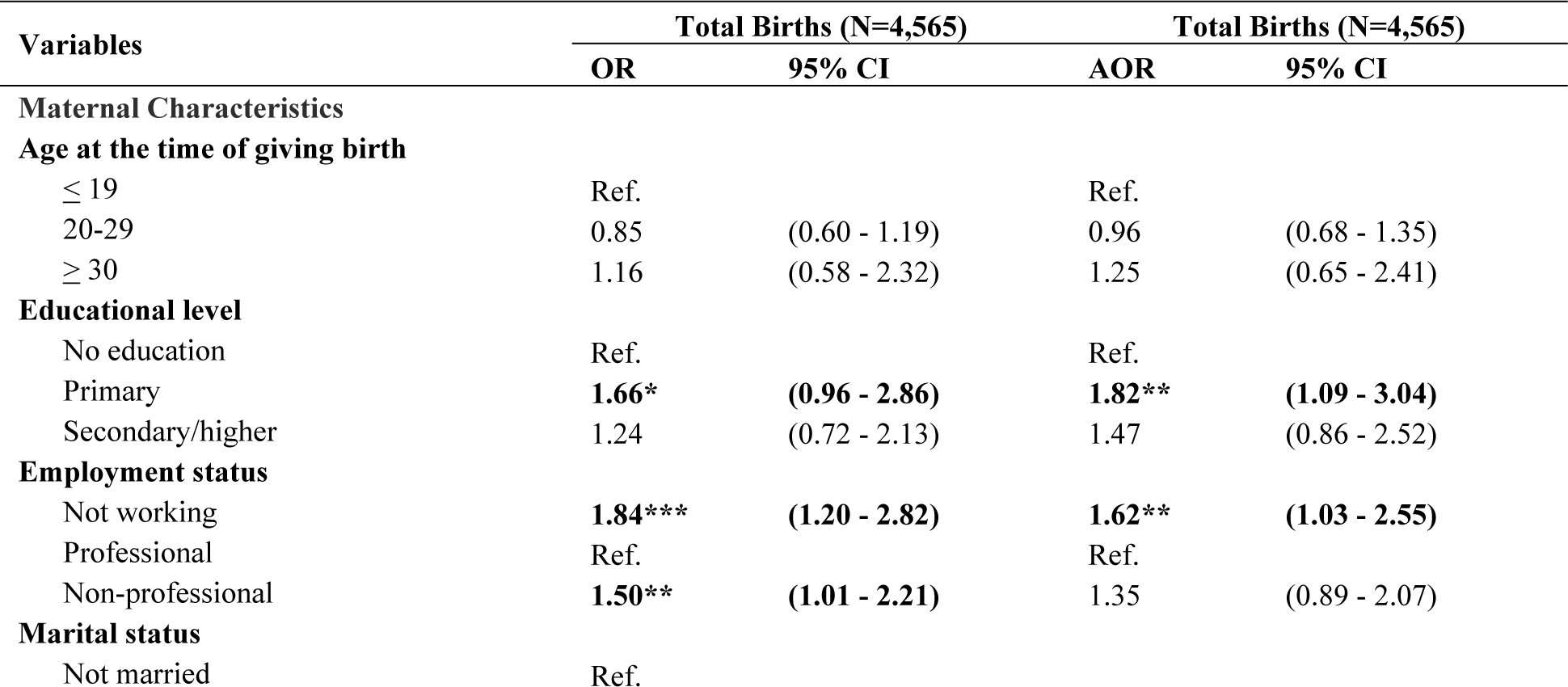
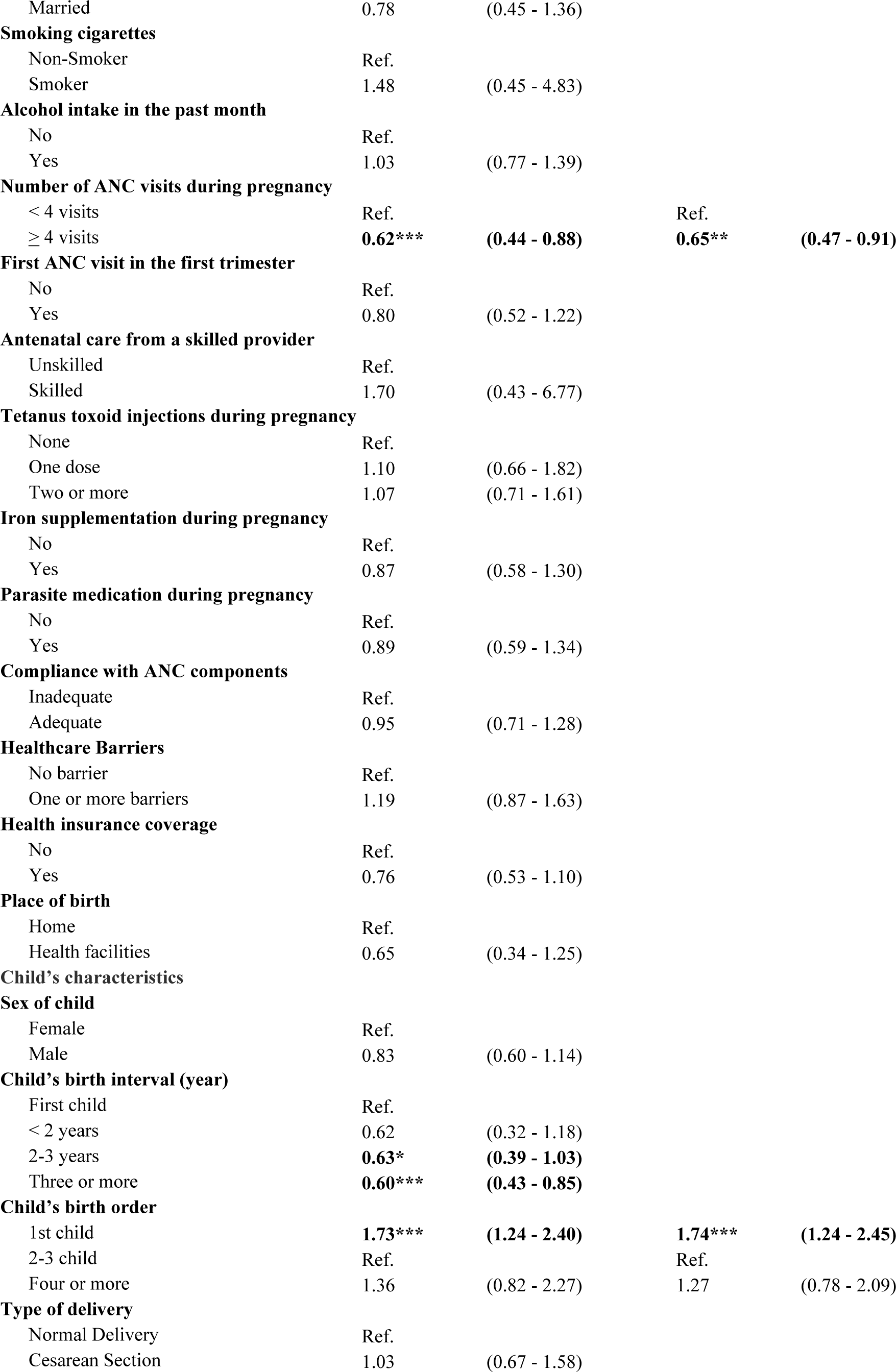
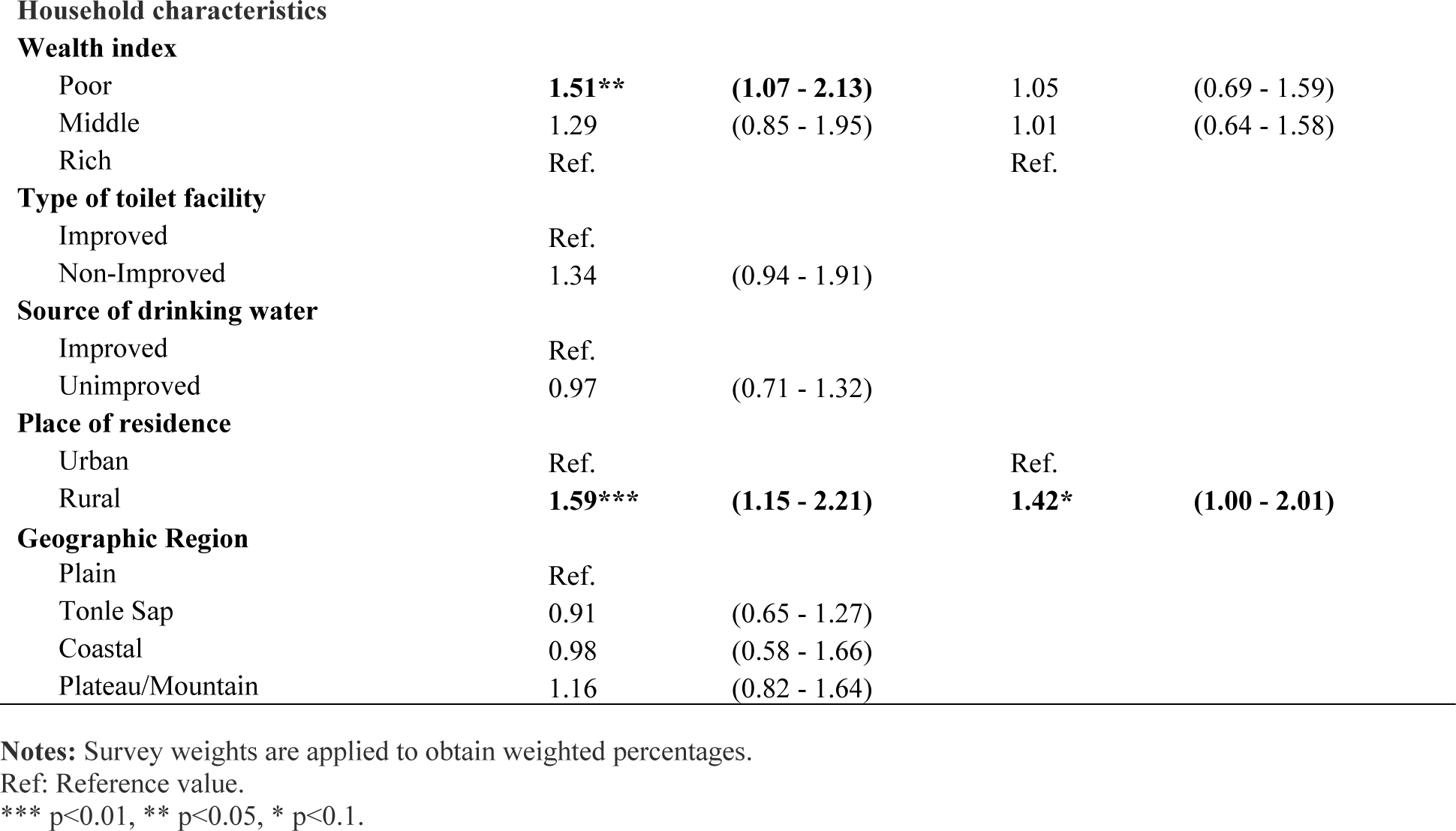
Factors associated with LBW among the most recent live births in Cambodia, CDHS 2021-2022.

| Variables | Total Births (N=4,565) |  | Total Births (N=4,565) |  |
| --- | --- | --- | --- | --- |
|  | OR | 95% CI | AOR | 95% CI |
| **Maternal Characteristics** |  |  |  |  |
| **Age at the time of giving birth** |  |  |  |  |
| ≤ 19 | Ref. |  | Ref. |  |
| 20-29 | 0.85 | (0.60 - 1.19) | 0.96 | (0.68 - 1.35) |
| ≥ 30 | 1.16 | (0.58 - 2.32) | 1.25 | (0.65 - 2.41) |
| **Educational level** |  |  |  |  |
| No education | Ref. |  | Ref. |  |
| Primary | **1.66\*** | **(0.96 - 2.86)** | **1.82\*\*** | **(1.09 - 3.04)** |
| Secondary/higher | 1.24 | (0.72 - 2.13) | 1.47 | (0.86 - 2.52) |
| **Employment status** |  |  |  |  |
| Not working | **1.84\*\*\*** | **(1.20 - 2.82)** | **1.62\*\*** | **(1.03 - 2.55)** |
| Professional | Ref. |  | Ref. |  |
| Non-professional | **1.50\*\*** | **(1.01 - 2.21)** | 1.35 | (0.89 - 2.07) |
| **Marital status** |  |  |  |  |
| Not married | Ref. |  |  |  |
| Married | 0.78 | (0.45 - 1.36) |  |  |
| **Smoking cigarettes** |  |  |  |  |
| Non-Smoker | Ref. |  |  |  |
| Smoker | 1.48 | (0.45 - 4.83) |  |  |
| **Alcohol intake in the past month** |  |  |  |  |
| No | Ref. |  |  |  |
| Yes | 1.03 | (0.77 - 1.39) |  |  |
| **Number of ANC visits during pregnancy** |  |  |  |  |
| < 4 visits | Ref. |  | Ref. |  |
| ≥ 4 visits | **0.62\*\*\*** | **(0.44 - 0.88)** | **0.65\*\*** | **(0.47 - 0.91)** |
| **First ANC visit in the first trimester** |  |  |  |  |
| No | Ref. |  |  |  |
| Yes | 0.80 | (0.52 - 1.22) |  |  |
| **Antenatal care from a skilled provider** |  |  |  |  |
| Unskilled | Ref. |  |  |  |
| Skilled | 1.70 | (0.43 - 6.77) |  |  |
| **Tetanus toxoid injections during pregnancy** |  |  |  |  |
| None | Ref. |  |  |  |
| One dose | 1.10 | (0.66 - 1.82) |  |  |
| Two or more | 1.07 | (0.71 - 1.61) |  |  |
| **Iron supplementation during pregnancy** |  |  |  |  |
| No | Ref. |  |  |  |
| Yes | 0.87 | (0.58 - 1.30) |  |  |
| **Parasite medication during pregnancy** |  |  |  |  |
| No | Ref. |  |  |  |
| Yes | 0.89 | (0.59 - 1.34) |  |  |
| **Compliance with ANC components** |  |  |  |  |
| Inadequate | Ref. |  |  |  |
| Adequate | 0.95 | (0.71 - 1.28) |  |  |
| **Healthcare Barriers** |  |  |  |  |
| No barrier | Ref. |  |  |  |
| One or more barriers | 1.19 | (0.87 - 1.63) |  |  |
| **Health insurance coverage** |  |  |  |  |
| No | Ref. |  |  |  |
| Yes | 0.76 | (0.53 - 1.10) |  |  |
| **Place of birth** |  |  |  |  |
| Home | Ref. |  |  |  |
| Health facilities | 0.65 | (0.34 - 1.25) |  |  |
| **Child's characteristics** |  |  |  |  |
| **Sex of child** |  |  |  |  |
| Female | Ref. |  |  |  |
| Male | 0.83 | (0.60 - 1.14) |  |  |
| **Child's birth interval (year)** |  |  |  |  |
| First child | Ref. |  |  |  |
| < 2 years | 0.62 | (0.32 - 1.18) |  |  |
| 2-3 years | **0.63\*** | **(0.39 - 1.03)** |  |  |
| Three or more | **0.60\*\*\*** | **(0.43 - 0.85)** |  |  |
| **Child's birth order** |  |  |  |  |
| 1st child | **1.73\*\*\*** | **(1.24 - 2.40)** | **1.74\*\*\*** | **(1.24 - 2.45)** |
| 2-3 child | Ref. |  | Ref. |  |
| Four or more | 1.36 | (0.82 - 2.27) | 1.27 | (0.78 - 2.09) |
| **Type of delivery** |  |  |  |  |
| Normal Delivery | Ref. |  |  |  |
| Cesarean Section | 1.03 | (0.67 - 1.58) |  |  |
| Wealth index |  |  |  |  |
| Poor | 1.51** | (1.07 - 2.13) | 1.05 | (0.69 - 1.59) |
| Middle | 1.29 | (0.85 - 1.95) | 1.01 | (0.64 - 1.58) |
| Rich | Ref. |  | Ref. |  |

Type of toilet facility
|  |  |  |
| --- | --- | --- |
| Improved | Ref. |  |
| Non-Improved | 1.34 | (0.94 - 1.91) |

Source of drinking water
|  |  |  |
| --- | --- | --- |
| Improved | Ref. |  |
| Unimproved | 0.97 | (0.71 - 1.32) |
| Place of residence |  |  |  |  |
| Urban | Ref. |  | Ref. |  |
| Rural | 1.59*** | (1.15 - 2.21) | 1.42* | (1.00 - 2.01) |

Geographic Region
|  |  |  |
| --- | --- | --- |
| Plain | Ref. |  |
| Tonle Sap | 0.91 | (0.65 - 1.27) |
| Coastal | 0.98 | (0.58 - 1.66) |
| Plateau/Mountain | 1.16 | (0.82 - 1.64) |

Notes: Survey weights are applied to obtain weighted percentages.
Ref: Reference value.
\*\*\* p<0.01, \*\* p<0.05, \* p<0.1.

## Discussion

The overall prevalence of LBW among newborns in Cambodia was 5.9% (95% CI: 5.2–6.8), which was lower than the percentage of LBW newborns that was 11% in 2000, 8% in 2005, and 7.9% in 2010 and 2014 [5,6,30], and lower than to the prevalence of LBW among newborns in South Asia was 24.9%, and global was 14% in 2020 [31]. The reduction in the prevalence of LBW corresponded with Cambodia’s efforts to reduce the mortality of neonates, infants, and those under five [7]. It might be attributed to the efforts and initiative of the Royal Government of Cambodia (RGC), which has strengthened health facilities across the country, particularly in rural areas, improved infrastructure, provided essential medical equipment and supplies, increased the number of midwives, expanded quality of antenatal care during pregnancy from a skilled provider, and provided more skilled medical practitioners at childbirth to ensure safe delivery practices at health facilities [24,32,33]. Moreover, the achievement can be attributed to the country’s concerted effort to increase women’s access to maternal health services [34]. Recent CDHS 2021-2022 indicated that institutional births dramatically increased, from 19.3% to 98%, had four or more ANC visits increased, from 9% to 86.1%, and had their first ANC visit during the first trimester also significantly increased from 10% to 87% between 2000 and 2022 [7]. In 2022, 98% of women took iron tablets or syrup, while 84% took intestinal parasite medications [7]. Recently, to encourage early and routine ANC visits, the government has offered pregnant women a monetary incentive of US$20 for each visit during a maximum of four ANC visits at any health facility with a contract with the National Social Security Fund (NSSF) [35,36].

The variation in the prevalence of LBW among newborns is highest in low-economic provinces such as Ratanak Kiri, Kratie, Pailin, Svay Rieng, and Stung Treng (see **Fig 1**). This may be due, at least in part, to limited implementation and enforcement of these policies and interventions in remote areas of the country [14].

Our study found several factors associated with increased odds of having LBW among newborns in Cambodia, including being born to mothers with primary education, being unemployed, being a first-born child, and being born to mothers in rural areas. However, mothers who attended at least four ANC visits during their last pregnancy were less likely to have LBW babies.

Mothers with primary schooling were 1.82 times more likely to have an LBW baby compared with those with a secondary or higher education, which is similar to the findings of 3 studies in other countries that the illiteracy of mothers is a risk factor for LBW [17,37,38]. This finding might be explained by the fact that highly educated mothers might have access to books and more education programs that help to better living conditions, knowledge of health processes such as antenatal care and nutrition, and access to quality healthcare services, all of which positively impact pregnant outcomes [9,25,26]. Similarly, unemployed mothers were 1.62 times more likely to have an LBW baby compared with those employed. This association is consistent with previous studies [39,40]. While a survey in several developing countries found that poverty is a risk factor for LBW [17,23,40,41], the present study found that these factors, although significantly associated with LBW in simple logistic regression analysis, were no relationship with LBW after controlling confounding factors in the multivariate logistic regression. Also, maternal age is considered a key factor for LBW [23,40]. This study reveals no statistical association between maternal age and low birth weight.

The study’s finding that mothers who received four or more ANC visits during their last pregnancy were 0.65 times less likely to have babies with LBW, compared with mothers who attended fewer than 4 ANC visits, is also in line with other studies elsewhere [17,42]. Therefore, the finding supports Cambodia’s MoH and WHO’s current national policy, which recommends at least 4 ANC visits during pregnancy [33]. Mothers with at least 4 ANC visits may receive proper ANC components such as weight monitoring, blood pressure measurement, blood sample collected, urine sample collected, nutrition counseling, ultrasound conducted, received counseling on danger signs, received two tetanus shots, received preventive deworming, received food supplements, and took iron and folic acid pills during the pregnancy [43]. This finding that the first child is more likely to have LBW compared with the second or third child is also in line with studies conducted in Cambodia by Chhea et al. 2018 [17]. The study found that children born to mothers’ residences in rural were 1.42 times more likely to have low birth weight babies. This association is consistent with previous studies [44]. A study in several developing countries found that rural residence is a risk factor for LBW [41]. This may be due, in part, to the limited implementation and enforcement of these policies and interventions in remote areas of the country. Rural areas have a higher poverty rate and restricted access to antenatal care services than urban areas [45,46].

### Strengths and limitations

Our study has serval strengths. First, this study used the most recent data from a nationally representative households survey with a high response rate of 97%. Data were collected using validated survey methods and highly trained data collectors, contributing to improved data quality. Second, the analysis was restricted to the last newborns with the most recent live births in the two years preceding the survey to explore variations of the LBW outcome across provinces, using recorded or recalled birthweight. This study’s findings contributed to the global prior prevalence of LBW and theoretical factors associated with LBW among newborns in Cambodia. Third, incorporating the DHS complex survey design and sampling weights into the analysis, it checked multicollinearity between independent variables and used a multiple logistic regression model to adjust for significant variables that may confound the association with LBW.

However, this study has several significant limitations. First, because CDHS 2021-2022 was collected as a cross-sectional nature of the data, the study is limited to associations of variables and not causality; the available data was obtained during the household survey, not during pregnancy or at the time of birth. While many variables, such as maternal age at birth, household wealth index, and place of residence, are unlikely to have changed, others, such as maternal employment status and marital status, may have changed between pregnancy or delivery and the survey. Another issue was dealing with missing data on the outcome variable for LBW. However, the problem is minor because less than 1% of the data was missing (CDHS 2021–2022). Second, it was restricted to independent variables for which CDHS 2021–2022 collected data. Many potential risk factors or contributing factors to low birth weight (LBW) were not well-documented. These included maternal malnutritional status (including anemia and weight gain during pregnancy), HIV infection, young married age, low birth weight, history of preterm births, and morbidity and illnesses during pregnancy, including non-communicable diseases like hypertension and diabetes.

### Conclusions

The findings suggest the overall prevalence of LBW is slightly decreased compared to CDHS 2014 and marginally lower compared to the Southsea Asia region. However, provincial variations in the prevalence of LBW were high in remote areas, and independent factors associated with LBW were identified. Interventions to reduce LBW should target provinces with relatively high prevalence, such as Ratanak Kiri, Kratie, Pailin, and Svay Rieng. Furthermore, the study shows that the mother’s educational level, unemployment, number of ANC visits during the last pregnancy, child’s birth order, and rural residence are significantly associated with LBW. These results strongly support and reconfirm Cambodia’s current national program policy that recommends pregnant women attend four or more ANC appointments during pregnancy. This policy should be further reinforced and implemented, considering finding effective ways to introduce pregnant who have limited education to promoting the quality of antenatal care among women in rural settings.

## Data Availability

Our study used the 2021-2022 Cambodia Demographic and Health Survey (CDHS) datasets. The DHS data are publicly available from the website at (URL:https://www.dhsprogram.com/data/available-datasets.cfm).

## Acknowledgments

The authors are grateful to DHS-ICF for gathering, cleaning, and providing the data used for this paper. We thank Dr. **Phorn Sreypheak** from the Khema Clinic in Phnom Penh, Cambodia, for her feedback and support.

## Abbreviations

ANC; Antenatal care, AOR; Adjusted odds ratio, CDHS; Cambodia Demographic Health Survey, EA; Enumeration areas, IFA; Iron folic acid, LBW; Low birth weight, NCDs; Non-communicable diseases, NECHR; National Ethical Committee for Health Research, NSSF; National Social Security Fund, RGC, Royal Government of Cambodia, PPS; Probability proportional to size, WHO; World Health Organization.

## Supporting information

**S1 Table.** Prevalence of LBW among newborns, CDHS 2021-2022 (n= 4,565 weighted count)

